# Therapeutic effectiveness of interferon-alpha 2b against COVID-19: the Cuban experience

**DOI:** 10.1101/2020.05.29.20109199

**Authors:** R Pereda, D González, HB Rivero, JC Rivero, A Pérez, LR López, N Mezquia, R Venegas, JR Betancourt, RE Domínguez

## Abstract

**Background:** Effective therapies are needed to control the SARS-Cov-2 infection pandemic and reduce mortality associated with COVID-19. Several clinical studies have provided evidence for the antiviral effects of type I interferons (IFNs) in patients with respiratory coronaviruses. This study assessed the therapeutic efficacy of IFN-α2b in patients infected with SARS-CoV-2 during the first month after the outbreak began in Cuba.

**Method:** This multicenter prospective observational study was conducted in 16 hospitals in 8 Cuban provinces. Participants were patients with confirmed SARS-CoV-2 infection detected from throat swab specimens by real time RT-PCR who gave informed consent and had no contraindications for IFN treatment. Patients received therapy as per the Cuban COVID protocol, that included a combination of oral antivirals (lopinavir/ritonavir and chloroquine) with intramuscular administration of IFN-α2b (Heberon^®^ Alpha R, Center for Genetic Engineering and Biotechnology, Havana), 3 times per week, for 2 weeks. The primary endpoint was the proportion of patients discharged from hospital (without clinical and radiological symptoms and non-detectable virus by RT-PCR). The secondary endpoint was the case fatality rate (CFR), defined as the number of confirmed deaths divided by the number of confirmed cases.

**Results:** From March 11^th^ to April 14^th^, 814 patients were confirmed SARS-CoV-2 positive in Cuba, 761 (93.4%) were treated with Heberon^®^ Alpha R and 53 received the approved protocol without IFN treatment. The proportion of fully recovered patients was higher in the IFN-treated compared with non-IFN treated group (95.4% vs 26.1%, *p*<0.01). The CFR for all patients was 2.95%, and for those patients who received IFN-α2b the CFR was reduce to 0.92. The estimated global CFR is 6.34% and 4.05% for the Americas reported by WHO and PAHO, respectively. In this study, 82 patients (10.1%) required intensive care and, of these, 42 (5.5%) were treated with IFN.

**Conclusions:** This report provides preliminary evidence for the therapeutic effectiveness of IFNα-2b for COVID-19 and suggests that the use of Heberon^®^ Alpha R may contribute to complete recovery.

## INTRODUCTION

The clinical spectrum of COVID-19 varies from asymptomatic infection to mild symptoms to severe acute respiratory illness and death^1,2^. There is an urgent need for antiviral drugs to effectively treat this disease.

Due to their antiviral properties and known mechanisms of action, type I interferons (IFN-α/β) present as candidate broad spectrum antivirals for global virus outbreaks^3^. In a recent clinical study, evidence was provided that IFN-α2b treatment accelerated viral clearance from the airways and reduced circulating levels of the inflammatory biomarkers IL-6 and CRP in COVID-19 cases^4^.

In previous coronavirus epidemics in 2002 (SARS-CoV) and 2012 (MERS-CoV), evidence was provided that coronaviruses encode in their genome factors that specifically block an IFN response, including preventing the activation of MyD88 associated with IFN production and STAT1, associated with IFN signaling^3^. Notably, for both outbreaks, evidence was provided for the antiviral effects of type I IFNs, suggesting that IFN treatment can override the inhibitory effects of coronaviruses^5, 6^.

In the absence of a vaccine, a number of candidate antivirals are currently under consideration around the globe. Remdesivir is considered the most promising ^7^; it functions by inhibiting the activity of RNA-dependent RNA polymerases (RdRp). Case studies describing the use of remdesivir for COVID-19 have been reported^8, 9^. Ongoing randomized, controlled clinical trials will evaluate the safety and antiviral efficacy of remdesivir in patients with mild to moderate or severe COVID-19 (NCT04292899, NCT04292730, NCT04257656, NCT04252664, NCT04280705).

Favipiravir, an anti-influenza medication that also is an RdRp inhibitor is under investigation for COVID-19, but to date, clinical data suggest limited efficacy^10^. The HIV protease inhibitor lopinavir/ritonavir (LPV/RTV) has seen disappointing outcomes in COVID-19 patients^11^, this despite evidence for LPV/RTV plus ribavirin being effective *in vitro* against SARS-CoV^12^.

The antiviral activities of chloroquine^13^ and hydroxychloroquine^14^ against SARS-Cov-2, prompted further evaluation in clinical studies, where early data suggest that they may contribute to inhibition of pneumonia exacerbation and shortening of disease course^15^.

Guidelines issued by the expert committee of the World Health Organization (WHO) identified IFN-α2b as a potential antiviral for the treatment and prevention of COVID-19^16^. Early on in the outbreak, the Chinese government recommended the use of IFN-α for the treatment of COVID-19^17^. Several ongoing clinical studies evaluating IFN-α2b for the treatment of COVID-19 are registered at clinicaltrials.gov^18^.

Heberon^®^ Alpha R (human recombinant IFN-α2b) produced by the Center for Genetic Engineering and Biotechnology, Havana, Cuba, has demonstrated antiviral efficacy and a proven safety profile over 34 years^19^. Herein, we report the first results of the use of Heberon® Alpha R as part of the Cuban protocol^20^ for the management of COVID-19.

## MATERIALS AND METHODS

### Patients and treatments

A multicenter, prospective, observational study was conducted, that included all patients in Cuba with confirmed SARS-CoV-2 infection during the first 33 days of the epidemic in the country, March 11 to April 14, 2020. Patients were recruited from 16 hospitals in 8 Cuban provinces. Individuals were considered COVID-19 positive based on virus detected by real time polymerase chain reaction (RT-PCR).

Patients with confirmed SARS-CoV-2 infection, who gave informed consent and had no contraindications for IFN treatment described in the product information sheet, received therapy as approved in the Cuban COVID protocol. Namely, a combination of antivirals and intramuscular Heberon^®^ Alpha R (IFN-α2b, liquid formulation) administration. Antivirals used were lopinavir/ritonavir (Kaletra) (250 mg, one capsule b.i.d. (500 mg/day) for 30 days and chloroquine 150 mg, one tablet twice a day (300 mg/day) for 10 days. IFN treatment was administered by intramuscular injection 3 million IU 3 times per week, for 2 weeks. To treat pediatric cases, the three drugs were adjusted for age and weight or body surface (i.e. IFN: 100.000 IU/Kg). Patients with contraindications or who did not consent to receive IFN were treated with the Cuban protocol lacking IFN, i.e. only lopinavir/ritonavir and chloroquine. For those cases whose disease progressed to become severe and critical, requiring ICU support, treatment with Heberon^®^ Alpha R was stopped.

Addition of Heberon^®^ Alpha R to the Cuban protocol was approved by the Cuban National Regulatory Authority, CECMED, which maintained surveillance of the scientific evidence obtained. The protocol of this study was evaluated and approved by a centralized Research Ethics Committee, representing all hospital institutions enrolled in the diagnosis and treatment of patients. This clinical research is registered with the code RPCEC00000318 in the Cuban Public Registry of Clinical Trials^21^.

Data presented are from patient medical records provided by the Cuban Ministry of Public Health, in accordance with the policies established for an investigation in a pandemic scenario.

### PCR confirmation of SARS-CoV-2

A qualitative real time polymerase chain reaction (RT-PCR) for SARS-CoV-2 was performed. Throat swab specimens from the upper respiratory tract of patients were placed into collection tubes prefilled with 150 μL of virus preservation solution, and total RNA was extracted using commercial kits: LightMix® Modular Sarbecovirus E-gene (Roche), LightMix® Modular SARS-COV-2 (COVID19 RDRP-GENE) (Roche), LightMix® Modular EAV RNA Extraction Control (Roche), LightCycler Multiplex RNA Virus Master (Roche), QIAamp® Viral RNA MiniKit (250) (Quiagen). RT-PCR positivity was required prior to treatment onset, and PCR testing was performed every 72 hours thereafter. Samples were designated positive (+) or negative (-).

Hematological and biochemical profiles were assessed at admission and every 72 hours using routine clinical laboratory procedures.

### Endpoints

The primary endpoint was the proportion of patients discharged from hospital (i.e. discharge criteria were the absence of clinical and radiological symptoms and non-detectable virus, as determined by 2 consecutive PCR(-)s at least 24 hours apart. The secondary endpoint was the case fatality rate, (CFR), defined as the number of confirmed deaths divided by the number of confirmed cases.

### Statistical analyses

Descriptive statistics were used to estimate the measures of central tendency and dispersion (media and extreme values) in demographic characteristics. The association between qualitative variables was analyzed using contingency tables. For the comparison of proportions the Fisher exact test calculator was used. The odds ratio (OR) was applied as a measure of association between treatments and outcomes. The non-parametric Mann-Whitney U test was used for comparing independents samples.

## RESULTS AND DISCUSSION

All confirmed patients with COVID-19 in Cuba during the period March 11^th^ to April 14^th^ 2020 were included in this study. The Cuban protocol^20^ for clinical management of COVID-19 was initiated on March 11^th^, 2020, concomitant with the diagnosis of the first three patients in the country. By April 14^th^, according to the Cuban Ministry of Public Health^22^, 814 individuals had been confirmed positive for SARS-CoV-2 infection, 761 of them (93.4%) were treated with the established protocol including Heberon^®^ Alpha R and 53 (6.6%) received the Cuban protocol without IFN.

**Table 1** describes the demographics of all patients included in this study. The distribution conformed to findings around the globe. The average age of all cases was 44.3 years, with a non-significant difference between the sexes.

**Table 1.** Demographics & clinical characteristics of patient cohort.

|  | IFN<br>n=761 | No IFN<br>n=53 | p value |
| --- | --- | --- | --- |
| Age<sup>a</sup> (years) | 42.9 (2-96) | 66.9 (1-101) | <0.01 |
| Male | 380 (49.9%) | 27 (51.1%) | 1.0 |
| Female | 381 (50.1%) | 26 (49.9%) | 1.0 |
| ICU<sup>b</sup> | 42 (5.5%) | 40 (75.5%) | <10<sup>-5</sup> |
| Co-morbidities<sup>c</sup> | 24 (3.2%) | 30 (56.6%) | <10<sup>-5</sup> |

<sup>a</sup> median and range are reported
<sup>b</sup> criteria for ICU: Required artificial ventilation, Acute Respiratory Distress Syndrome.
<sup>c</sup> high blood pressure, diabetes mellitus, ischemic heart disease, chronic kidney failure, asthma, obesity, chronic obstructive pulmonary disease, pancreatic cancer, liver cirrhosis, bladder neoplasia, gastritis, hypothyroidism, glaucoma, myasthenia gravis, HIV.

Patients who did not receive IFN were older (median 66.9 years vs 42.9 years) with a higher incidence of comorbidities (56.6% vs 3.2%), such as high blood pressure, ischemic heart disease and diabetes mellitus.

Data analyses were limited, because the study endpoint outcomes, namely hospital discharge (recovery) and CFR for a large number of the cases remained unknown at the time of study termination. On April 14^th^, 639 patients remained hospitalized under protocol care (herein considered as patients with unknown outcome).

For individuals with known outcomes (**Table 2**), a higher proportion of patients who became PCR SARS-CoV-2 negative and resolved their disease, were IFN-treated, than not (95.4% vs 26.1%, *p*<0.01). According to the Odds Ratio estimation, an individual treated with Heberon^®^ Alpha R had a 58.7 times greater advantage to achieve recovery.

**Table 2.**
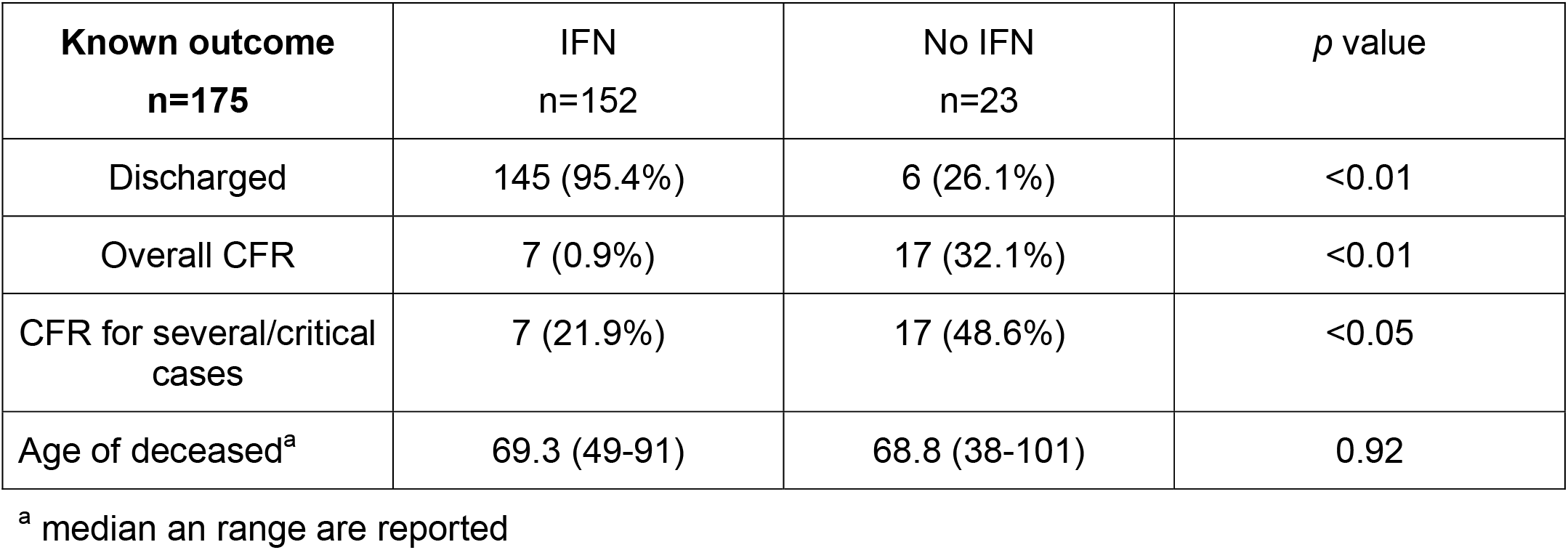
IFN treatment promotes recovery from COVID-19.

On the cut-off date (April 14, 2020), the overall Cuban CFR was 2.95%^22^. The CFR for patients treated with IFN-α2b was 0.92 (*p*<0.01). These CFRs are lower than those reported on the same date by the WHO^23^: Global CFR=6.34% and PAHO^24^ CFR=4.05% (for the region of the Americas).

To better address the influence of IFN treatment on CFR, we next considered the clinical status of patients in the context of severity of disease, using admission to ICU as a criterion for severe disease.

82 patients (10.1% of all the COVID-19 cases) required intensive care, of which 42 had been treated with IFN. Notably, this is 5.5% of the total number of COVID-19 cases that had been treated with IFN. 67 (8.2% of all the COVID-19 cases) patients progressed to a serious or critical state of the disease, as characterized by a requirement for artificial ventilation and Acute Respiratory Distress Syndrome. 35 of these had not been treated with IFN-α2b (representing 66.0% of COVID-19 cases that had not received IFN treatment). By contrast, 32 patients who progressed to serious or critical disease had received treatment with IFN-α2b (representing 4.2% of the COVID-19 cases that had received IFN treatment). The data suggest that treatment with IFN limits progression to severe disease *(p<0.05)*. IFN treatment was withdrawn for those patients who progressed to critical disease. Nevertheless, the effects of IFN treatment on subsequent CFR revealed that prior IFN treatment was associated with a 3.37 times greater likelihood of survival, since those critical patients not treated with IFN (48.6%) did not survive, compared to 21.9% of the IFN-treated patient who did survive (*p*<0.05).

In this study the median age was higher in those patients that did not receive IFN. Despite this, data analysis revealed that age was not a variable that negated the effects of IFN treatment.

Notably, the CFR of COVID-19 by age was consistent with data from other countries that suggest that the elderly are at greater risk for severe disease and death^13^. In our study, 22 of the 24 (91.7%) fatalities, were older than 44.3 years and 19 (79.2%) were older than 55 years.

There were more deaths among males: 16 males compared with 8 females. Our data indicate that the estimated CFR in males with comorbidities is higher for individuals above the age of 45 years: 16.5 times greater likelihood of death.

Irrespective of age, CFRs for those with comorbidities is much higher (87.5% vs 0.52%). In our study cohort, 7 (1.8%) of those under 45 years of age and 47 (11.4%) older had co-morbidities. Those individuals with comorbidities died: 21 (38.9%), compared to those with no co-morbidity 3 (0.39%). Elderly age and arterial hypertension represented the major risk predictors for death; in patients over 55 years, this increased from 4.9% to 38.9%.

The lower incidence of comorbidities in the IFN-treated patients may have contributed to their 4.34-fold greater survival advantage compared with those had not been treated with IFN (**Table 1**). However, this advantage disappears in the subgroup of 28 patients with high blood pressure. High blood pressure was a comorbidity in 27 (32.9%) of the 82 patients who required intensive care. After adjusting for demographic characteristics, the IFN-related CFR was affected by comorbidity (*p*=1 × 10^-4^) and age (*p*=0.02) and remained significant regardless of sex (*p*=0.29).

The limitations of this open, non-randomized observational study include unbalanced demographics between treatment arms of unequal size. Nevertheless, the purpose of this study was to rapidly evaluate if inclusion of IFN-α2b at the doses and therapeutic regimen employed, offered a therapeutic benefit to COVID-19 cases. Recent publications suggest that treatment with chloroquine^16^ or lopinavir/ritonavir^11^ may offer little therapeutic benefit in COVID-19. With this information, we postulate that IFN treatment is likely the active antiviral in the regimen employed. Regardless of the identified limitations, therefore, our findings suggest that IFN-α2b treatment may be effective for the treatment of COVID-19

## CONCLUSION

This report provides evidence of the effectiveness of IFN-α2b as an antiviral treatment for COVID-19 and suggests that the use of Heberon^®^ Alpha R may contribute to recovery from COVID-19. These data extracted from clinical records of all patients in the first month of the epidemic, reveals that the use of Heberon^®^ Alpha R improved both the rates of recovery and case fatalities.

## Data Availability

Here I state the availability of all data referred to in the manuscript

## Contributors

DG, RV, BJR and DRE were responsible for patient care and treatment, RHB, RJC, OA, LLR and MN made clinical oversight and clinical data collection, PR led the working group, analyzed and conducted data analysis, data interpretation, literature searches and manuscript writing.

## Acknowledgement

The authors are grateful to Dr. Eleanor Fish from Toronto General Hospital Research Institute, University Health Network and Department of Immunology, University of Toronto, Toronto, ON, Canada, who reviewed the data and assisted with the interpretation and writing of the manuscript.

